# Research involvement of medical students in a medical school of India: exploring knowledge, attitude, practices, and perceived barriers

**DOI:** 10.1101/2024.05.19.24307598

**Authors:** Abhinav Jha, Manas Ratan Shah, Ritik Goyal, Deepak Dhamnetiya, Apoorv Amitabh Bharatwal, Ravi Prakash Jha, Prachi Renjhen

**Affiliations:** Dr Baba Saheb Ambedkar Medical College and Hospital, Rohini Sector 6, New Delhi-110085; Department of Community Medicine, Atal Bihari Vajpayee Institute of Medical Sciences and Dr. Ram Manohar Lohia Hospital, Baba Kharak Singh Marg, Connaught Place, New Delhi India-110001; Department of Community Medicine, Dr Baba Saheb Ambedkar Medical College and Hospital, Rohini Sector 6, New Delhi-110085; Department of Obstetrics and Gynaecology, Dr Baba Saheb Ambedkar Medical College and Hospital, Rohini Sector 6, New Delhi-110085

**Keywords:** Research, Medical Students, Knowledge, Attitude and Practices, Research Society, Medical School

## Abstract

**Introduction:** Research in the medical discipline significantly impacts society by improving the general well-being of the population, through improvements in diagnostic and treatment modalities. However, of 579 Indian medical colleges, 332 (57.3%) did not publish a single paper from the year 2005 to 2014," indicating a limited contribution from medical fraternity In order to probe in to the cause of this a study was conducted to assess the knowledge, attitude, practices (KAP) and perceived barriers to research among students of a medical school in Delhi, India.

**Methods:** A cross-sectional study was conducted among medical students and the data on academic-cum-demographic information, assessment of knowledge, attitude, practices and barriers to research was collected using a pre-tested, semi-structured questionnaire. Chi-square test was used to check the association of various factors with the KAP of research. A p-value less than 0.05 was considered significant.

**Results:** A total of 402 (N) subjects were enrolled in the study. Majority were male (79.6%) and from clinical professional years (57%). Majority (266, 66.2%) of the subjects had adequate knowledge. Of the study subjects (61,15%) having inadequate knowledge of research, sixty percent were from pre- and para-clinical years, while around 70 % of those having good knowledge were from clinical professional years. However, only 16.9% of the participants had participated in a research project, and only 4.72% had authored a publication. Sixty one percent of study subjects having a positive attitude towards research, were from pre- and para-clinical years. Among the study subjects having a positive attitude towards research, over 60% were from pre- and para-clinical years. The barriers for conducting research were mostly; lack of funds/laboratory equipment/infrastructure (85.1%), lack of exposure to opportunities for research in the medical (MBBS) curriculum (83.8%), and lack of time (83.3%). There was a statistically significant association between knowledge and attitude towards research with a professional year of study.

**Conclusions:** The study revealed that while most of the students had a positive attitude towards research as well as an adequate knowledge of research, there was a poor level of participation in research. These challenges can be overcome by incorporating research as a part of the medical school curriculum from early years on, setting aside separate time for research, and establishing student research societies that can actively promote research.

## Introduction

The World Health Organisation (WHO) defines research as a quest for knowledge through diligent search, investigation, or experimentation to discover and interpret new knowledge (1). The purpose of which is to advance knowledge through the development of scientific theories, concepts and ideas with the ultimate goal of improving human lives and enhancing society. Research in the medical discipline bears tremendous significance in society by contributing to increased longevity of people, through improvements in diagnostic and treatment modalities, ultimately contributing to increased productivity and economic stability of the population. Conducting research holds significant importance for clinicians as it enables them to stay abreast of disease trends and associated risk factors. Moreover, it facilitates the examination of treatment outcomes and public health interventions, ultimately contributing to the delivery of higher-quality healthcare (2).

Physician-scientists play a crucial role in bridging the gap between basic and clinical sciences. They uncover the molecular basis of diseases and translate that knowledge into therapeutic interventions (3). As a medical student, conducting research is an important part of the medical school/college experience, as it fosters critical thinking and analytical skills through hands-on learning, defines an individual’s academic, career and personal interests, expands knowledge and understanding of a chosen field beyond the classroom and helps in building connections with peers, faculty and organizations on- and off-campus. The clear observation is that introducing medical students to clinical research activities at an early stage enhances their inclination toward pursuing an academic medical career. Even if these students do not opt for a path in clinical research, their understanding of research principles will enhance their capability to make evidence-based decisions in clinical practice (3). It has been shown that having a medical school publication positively influences academic career choice among medical students (4).

Despite India’s emergence as the world’s third-largest publisher of science and engineering articles, accounting for 5.31% of global research output with 135,788 published articles in 2018 (5), a significant portion of this research originates from faculty members or scientists affiliated with prestigious medical institutes. A review of research publications from 579 Indian medical colleges and hospitals revealed that "Out of 579 Indian medical colleges, 332 (57.3%) did not publish a single paper from the year 2005 to 2014," indicating a limited contribution from medical students as well as professors (6).

There is a need to address the gap in the medical education system of India which concentrates mainly on preparing doctors who are trained in allopathic sciences but seldom promotes research activities. Students in India have almost no formal pathway for becoming physician-scientists (7). With an evidently high Indian population and huge disease burden, there are enormous opportunities for doing research and exploring new solutions (8,9). Literature search reveals scarce data on knowledge, attitude and practices of research among medical students of Delhi, India. This study aims to assess the knowledge, attitude, practices, and perceived barriers towards research among students of a medical school in Delhi.

## Materials and Methods

To address the above-stated aims, a cross-sectional study was conducted during the month of May-June 2023 among the students of a medical college in Delhi, North India, after getting the approval from the Institutional Ethical Committee (IEC). All the medical students who were enrolled at our institute were included in the study after taking online written consent. The students who did not give consent or dropped out from our institute were excluded from this study. The required sample size for present study was determined using the formula 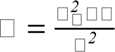. With a 95% confidence interval, a response distribution of 50%, and a margin of error of 0.05, the calculated minimum sample size of 384 participants was determined to adequately represent the population of medical students.

### Ethical Consideration and Confidentiality

Ethical approval for this study was provided by the institutional research review committee (IRRC) and institutional ethical committee (IEC) and online informed consent was taken from each participant. Participants were allowed to withdraw their names at any given time. The confidentiality of the data was ensured by maintaining the responses anonymous.

### Data collection

Data was collected using a pre-tested, semi-structured questionnaire. The proforma was pretested among 10% of the subjects. The questionnaire consisted of 5 sections, a total of 47 questions, Section 1 consisted of questions regarding demographic information like age, gender and academic-year of study. Section 2 consisted of 16 questions regarding knowledge of research, Section 3 consisted of 11 questions regarding attitude towards research, Section 4 consisted of 11 questions regarding the barriers to research and Section 5 consisted of 5 questions regarding practices of research. The final questionnaire was created using Google forms and data was collected using the same Google form link on the official WhatsApp (a messaging software app) group of our medical school (college). A total of 402 responses were collected and analysed for the final results.

### Data Management and Statistical Analysis

The collected data was coded and recorded in a Microsoft Excel sheet. Categorical data were analyzed using percentages and proportions, while quantitative data were summarized using the mean and standard deviation. The scoring for knowledge of research was used to put the participants into three categories: inadequate knowledge (score<5), adequate knowledge (scoring 5–10), and high knowledge (score > 10). In addition, participants were categorised into three categories according to their attitude towards research: negative attitude (scoring 0-3), neutral attitude (score 4-5), and positive attitude (score 6-8). Additionally, four questions about research practices were included. These were scored on a Likert scale from 1 to 5, with 1 denoting strongly disagree, 2 disagree, 3 neither agree nor disagree/neutral, 4 denoting agree, and 5 denoting strongly agree. Chi-square test was used to check the association of various factors with the KAP of research. The trial version of Statistical Package for the Social Sciences (version 27.0; SPSS Inc., Chicago, IL) was used for the analysis of the data. A p-value below 0.05 was considered significant.

## Results

A total of 402 (N) subjects were enrolled in our study. Majority (320,79.6%) of the subjects were male and belonged to the 18-21 age group (262,65.2%). We had more participants (229,57%) from the Clinical professional year. Most (266,66.2%) of the subjects had adequate knowledge of research, while few (61,15.2%) subjects had inadequate knowledge. Most (328,81.6%) of the subjects have a positive attitude towards research, while few (32,8%) have a negative attitude. Only 16.9% (68) had been a part of a research project, while only 4.72% (19) of the subjects had a publication. Only 6.96% (28) had presented their research in a Conference. Only 8.95% (36) of the subjects had attended a research methodology workshop (Table 1).

**Table 1:**
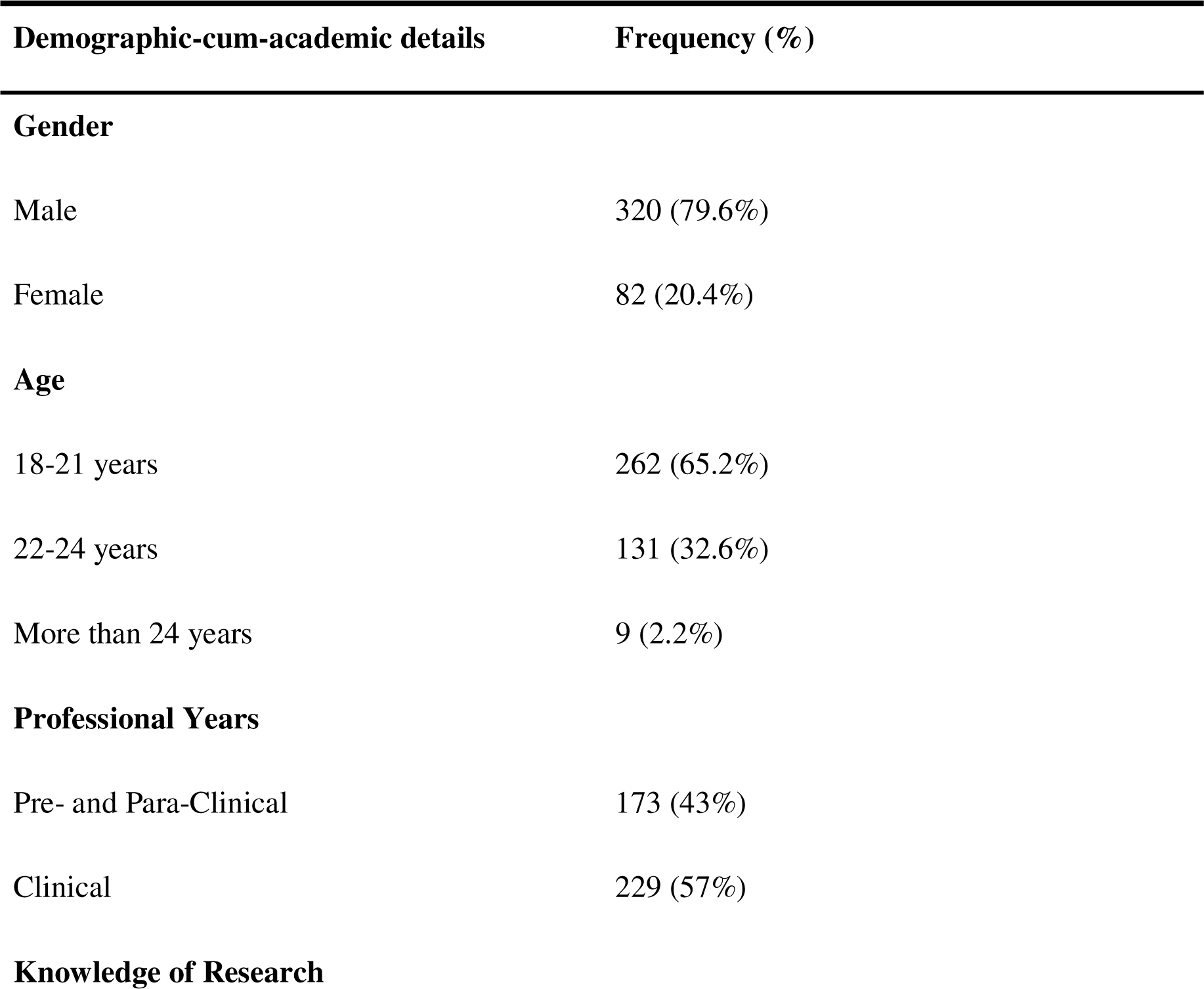
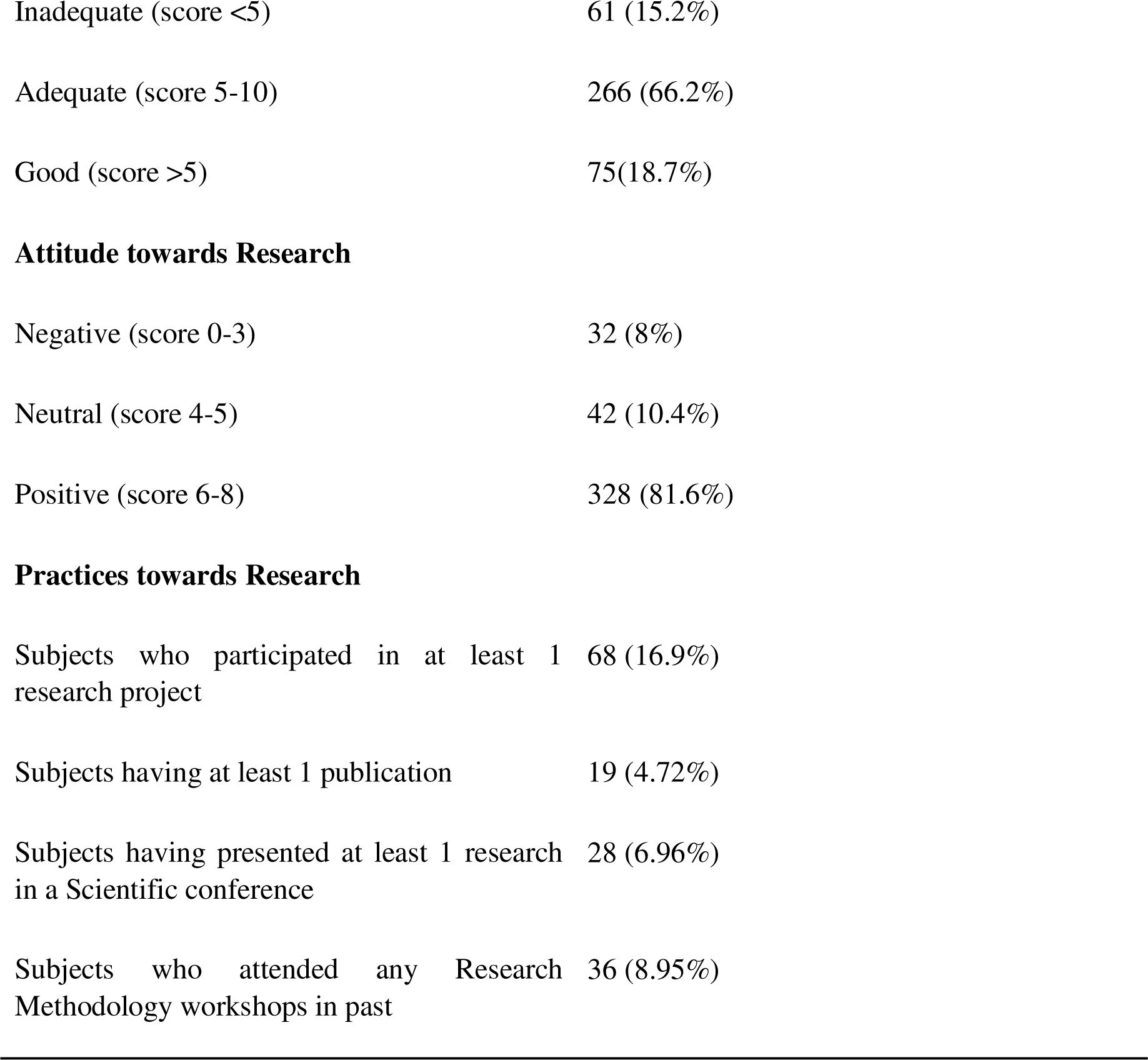
Demographic-cum-academic profile and KAP comprehensive score categories of the study subjects (N=402)

| Demographic-cum-academic details | Frequency (%) |
| --- | --- |
| **Gender** |  |
| Male | 320 (79.6%) |
| Female | 82 (20.4%) |
| **Age** |  |
| 18-21 years | 262 (65.2%) |
| 22-24 years | 131 (32.6%) |
| More than 24 years | 9 (2.2%) |
| **Professional Years** |  |
| Pre- and Para-Clinical | 173 (43%) |
| Clinical | 229 (57%) |
| **Knowledge of Research** |  |
| Inadequate (score <5) | 61 (15.2%) |
| Adequate (score 5-10) | 266 (66.2%) |
| Good (score >5) | 75(18.7%) |
| **Attitude towards Research** |  |
| Negative (score 0-3) | 32 (8%) |
| Neutral (score 4-5) | 42 (10.4%) |
| Positive (score 6-8) | 328 (81.6%) |
| **Practices towards Research** |  |
| Subjects who participated in at least 1 research project | 68 (16.9%) |
| Subjects having at least 1 publication | 19 (4.72%) |
| Subjects having presented at least 1 research in a Scientific conference | 28 (6.96%) |
| Subjects who attended any Research Methodology workshops in past | 36 (8.95%) |

There was no significant association between knowledge or attitude towards research and category of age or gender. We found a statistically significant association between knowledge of research and professional year. Among participants having inappropriate knowledge of research, 61% (37) were from pre and para clinical years. While among those having good knowledge of research, 69% (52) were from clinical years. We also found a statistically significant association between attitude towards research and professional year. Among the study subjects having a negative attitude towards research, 72% (23) were from clinical years. While among those having a positive attitude towards research, 61% (200) were from pre and para clinical years (Table 2).

**Table 2:** Association of age, gender and academic-professional year with knowledge and attitude towards research (N=402)

| Variable | Knowledge of Research |  |  | p- value |
| --- | --- | --- | --- | --- |
|  | Inadequate (n=61) | Adequate (n=266) | Good (n=75) |  |
| Age in years |  |  |  |  |
| 18-20 | 27 (44.26%) | 112 (42.10%) | 24 (32.0%) | 0.122 |
| 21-23 | 32 (52.45%) | 137 (51.50%) | 41 (54.66%) |  |
| ≥24 | 2 (3.27%) | 17 (6.39%) | 10 (13.33%) |  |
| Gender |  |  |  |  |
| Male | 52 (85.25%) | 208 (65.0%) | 60 (80.0%) | 0.466 |
| Female | 9 (14.74%) | 58 (70.7%) | 15 (20.0%) |  |
| Professional year |  |  |  |  |
| Pre- and para-Clinical | 37 (60.65%) | 169 (73.8%) | 23 (30.66%) | 0.000 |
| Clinical | 24 (39.35%) | 97 (56.1%) | 52 (69.33%) |  |
| Attitude towards Research |  |  |  |  |
|  | Negative (n=32) | Neutral (n=42) | Positive (n=328) | p- value |
| Age in years |  |  |  |  |
| 18-20 | 7 (21.87%) | 14 (33.33%) | 142 (43.29%) | 0.065 |
| 21-23 | 24 (75.0%) | 24 (57.14%) | 162 (49.38%) |  |
| ≥24 | 1 (3.12%) | 4 (16.66%) | 24 (7.31%) |  |
| **Gender** |  |  |  |  |
| Male | 26 (81.25%) | 33 (78.57%) | 261 (79.56%) | 0.960 |
| Female | 6 (18.75%) | 9 (21.42%) | 67 (20.42%) |  |
| **Professional year** |  |  |  |  |
| Pre- and para-Clinical | 9 (28.12%) | 20 (57.62%) | 200 (60.97%) | 0.001 |
| Clinical | 23 (71.88%) | 22 (42.38%) | 128 (39.02%) |  |

Most (361, 89.8%) of the study subjects found research interesting and 79% (319) wanted to participate in a research methodology workshop. Eight-eight percent (353) of students wanted the teaching of research methodology to be a part of the MBBS curriculum. and sixty percent (242) of the subjects wanted research to be one of the criteria for acceptance in residency/post-graduate medical specialty programs in India (Figure 1). Among the study subjects, most of them agreed: willingness to conduct research (126,31.3%), willingness to spend time on a research internship (132,32.8%), conducting research even if it doesn’t lead to a publication (103,25.6%). 25% (100) of the subjects were neutral to devote the same time for research as for university exams (Figure 2).

**Figure 1:**
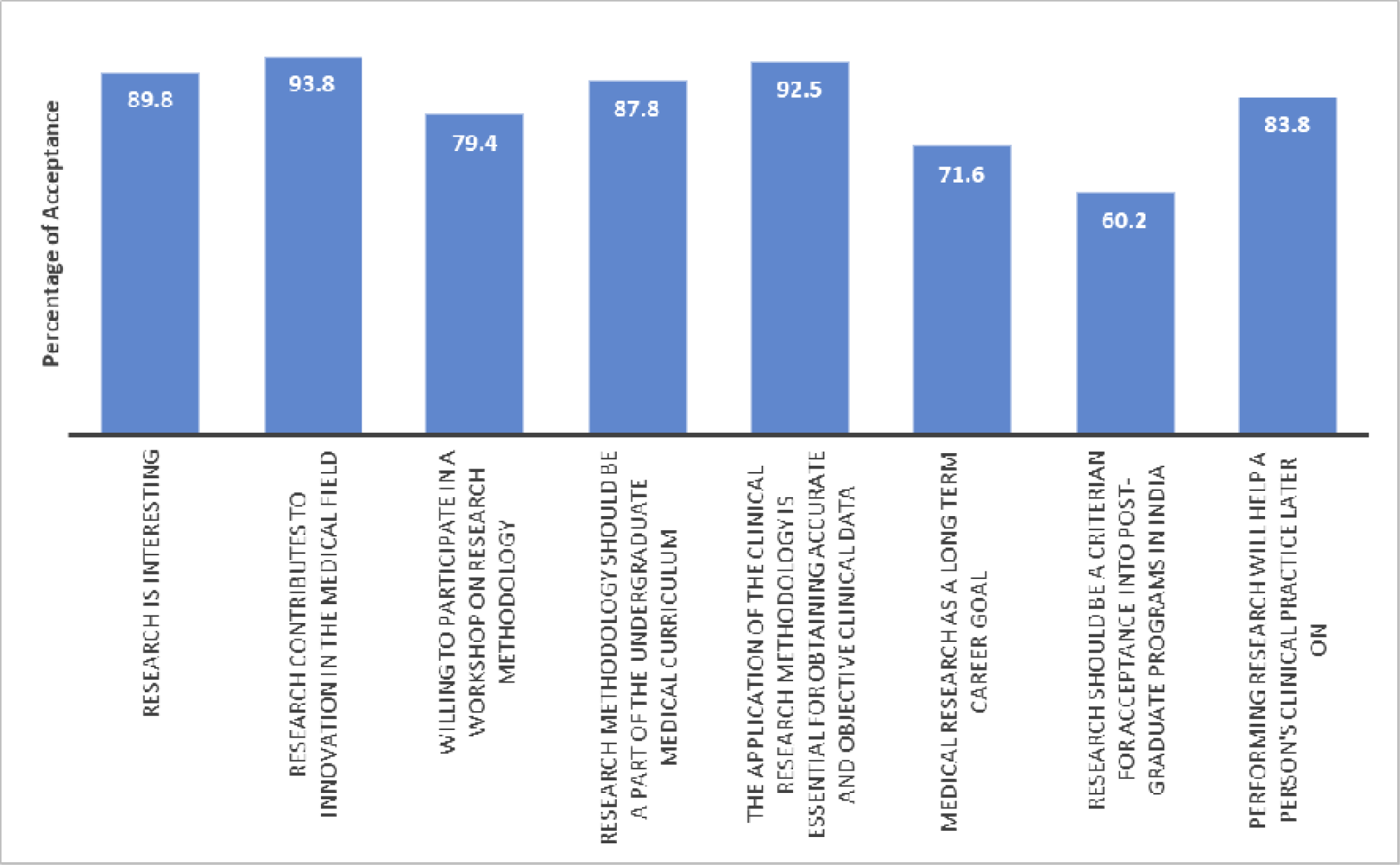
Response of study subjects to the questions assessing attitude towards research (N=402)

**Figure 2:**
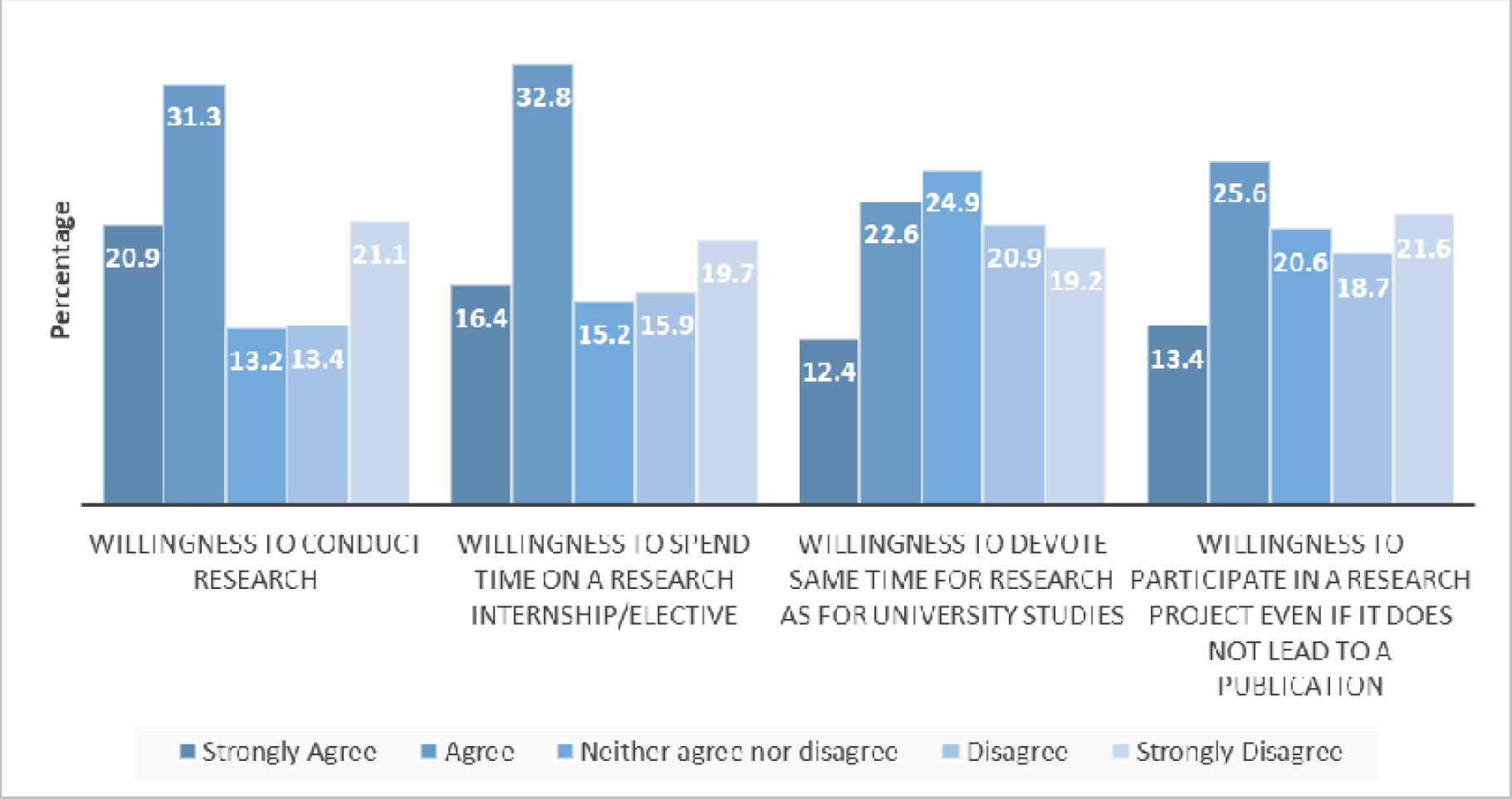
Distribution of the participants according to attitude regarding the following research practices (N=402)

The motivation for starting a research project were mostly their own interest (243,60.4%), to enhance academics (169,42%) or for attaining publications (160,39.8%). Only a few wanted to conduct research for financial benefits (85,21.1%) or because of peer pressure (27,6.7%) (Figure 3). According to the subjects, the barriers for conducting research were mostly: lack of funds/laboratory equipment/infrastructure (342,85.1%), lack of exposure of opportunities for research in MBBS curriculum (337,83.8%), lack of time (335,83.3%), difficulty in finding a supervisor (313,77.9%) and lack of incentives for conducting research (310,77.1%) (Figure 4).

**Figure 3:**
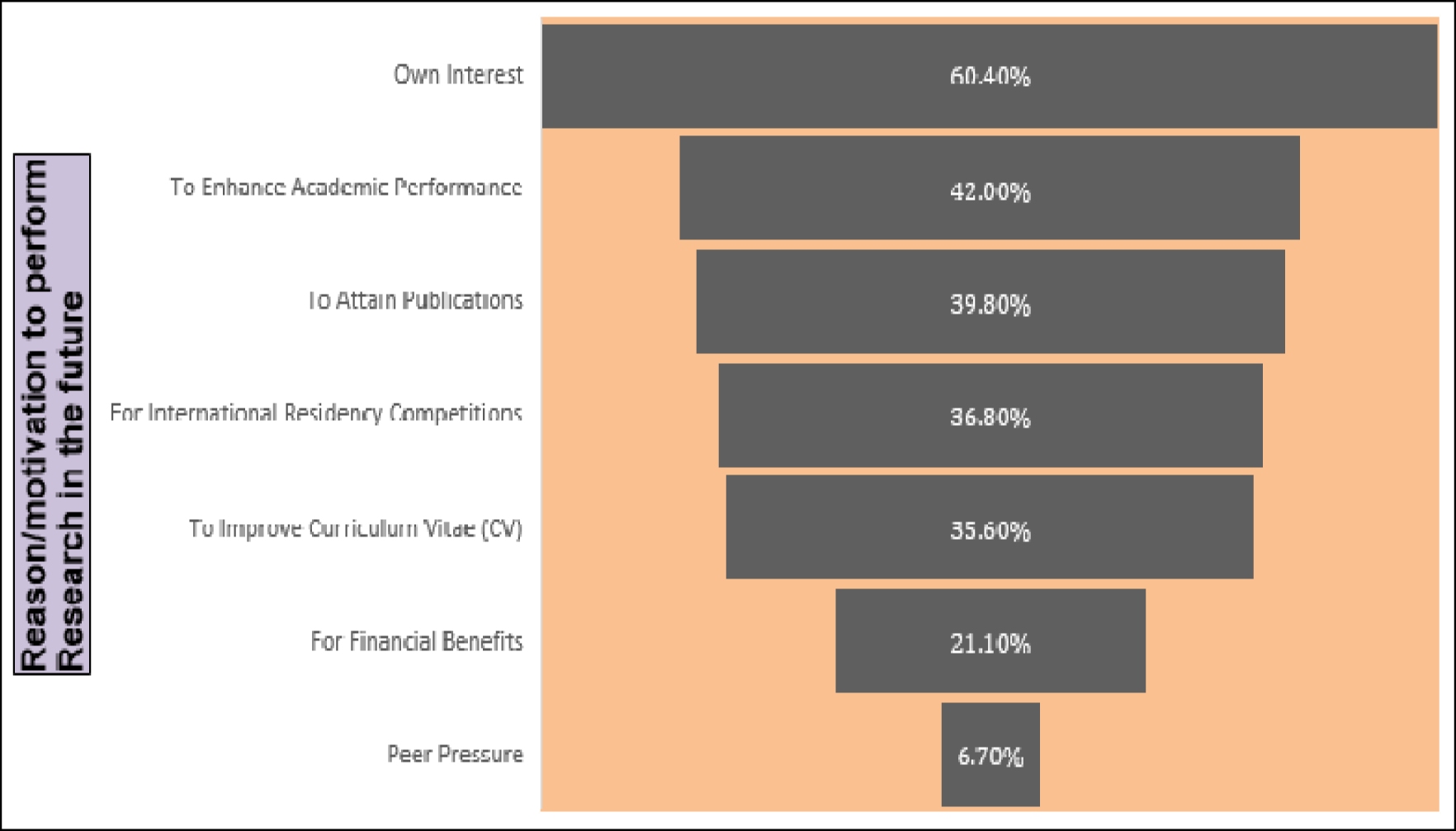
Reason/motivation to conduct research in the future (N=402)

**Figure 4:**
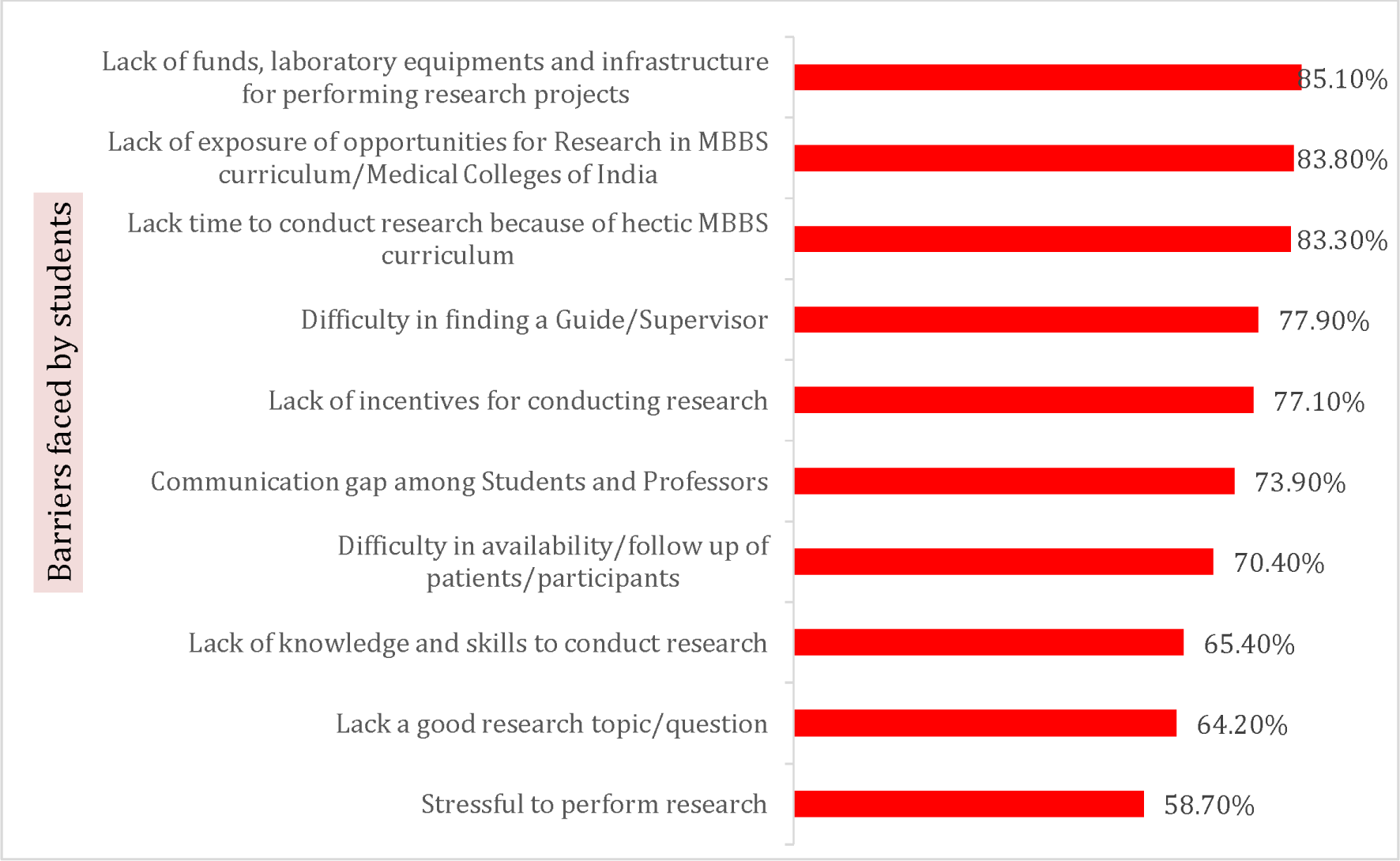
Barriers for conducting research (N=402)

## Discussion

We found that most of the participants had a positive attitude towards research. Most (89.8%) of the subjects found research to be interesting and believed that research contributed to innovations in the medical field, similar to a previous study (97%) conducted by Omprakash et al (11).

Most (92.5%) of the participants felt that performing research would help in their clinical practice in the future and most (71.6%) saw medical research as their long-term career goal, similar results also reported in studies by Abushouk et al., Ahmed et al., Vairamani et al., Ibrahim et al., Ismail et al. (3,12–15). Few research papers, though, produced contradictory findings such as Kumar et al. reported that only 6% of subjects were interested in research as a future career (16). Siemens et al. reported that only 44% of subjects agreed or strongly agreed that research will play a significant role in their future career (17). Meraj et al. reported that 70% students perceive research as stressful which 42% of the students did not see medical research as a long-term career goal (18) which can be due to the perceived bias against research as a non-lucrative career or the uncertainty on what a future in research might behold for them.

Nearly eighty eight percent of the study participants wanted research methodology to be a part of the curriculum and were willing to participate in such workshops. These findings were comparable with previously conducted studies by Omprakash et al. And Vairamani et al. where 84% and 76% of the students respectively wanted research to be the part of their curriculum (11,13). However, only 60.2% felt that research should be a criterion for acceptance in postgraduate programs in India indicating that even though many participants wanted to enhance their knowledge about research, they were against mandating it. Similar results were documented by Ahmed et al. and Siemens et al. where 58% and 47% of participants respectively didn’t want research as mandatory criteria for admission in Post Graduation (12,17).

Almost half of the participants were willing (or strongly willing) to conduct research as well as in spending time on research internships/electives, similar to observations made by Ibrahim et al. In his study 61% participants wanted to conduct clinical research (3), however, only 40% of the participants were willing to devote the same time for research as their university studies. This signifies that while participants do want to devote some of their time towards research, academics was a precedent for them.

Most (66.2%) of the study participants had adequate knowledge (score range 5-10) about research. This is in contrast to previously conducted studies by Abushouk et al. and Amin et al. which revealed low knowledge scores with average scores of 2.4 out of 6 and 3.5 out of 10 among the participants (3,19). Ibrahim et al. also showed that 67% of students had low knowledge (14). while another study by Pallamparty et al. revealed 70% subjects have good knowledge (20). The contrasting results can be attributed to the varying difficulty of the questions.

Most of the participants were aware of the definition of research (97%), the requirements of the IEC (90.3%), bias (84.1%), citations and references (87.1%). However, only a few subjects (11.7%) were aware of the first step in a research study, and referencing software. This indicates that the participants were aware of some theoretical aspects of research which are covered in the curriculum being followed. However, there exists a knowledge gap about the practical aspects of research.

We found a statistically significant association between knowledge of research and professional year of study. Among the study subjects having inadequate knowledge of research, 61% were from pre- and para-clinical years. Among those having good knowledge of research, 69% were from clinical years. These results are similar to a previously conducted study by Khan et al. which postulated that with increase in one study year, knowledge increases by 4.1%(21). It can be postulated that as the medical students of India, in their initial phase of the clinical year (3rd year) undergo annual assessment in the subject of community medicine about some aspects of research like basics of epidemiology, study designs, biostatistics, etc., has led to them to score higher on the knowledge score than their fellow peers from pre- and para-clinical years. Hren et al. found that the maximum score was in year 3 students which could have been due to them finishing their mandatory course on the principles of scientific research in medicine (22).

There was a statistically significant association between attitude towards research and professional year. Participants from pre- and para-clinical years had a more positive attitude towards research compared to those from clinical years. One plausible rationale for this finding could be the increased academic burden because of the lengthy syllabus during the clinical years, coupled with the pressure of competitive exams for postgraduate residency. This is in contrast to a previous study conducted by Khan et al. (21) which found, that the participants’ knowledge and attitude towards health research improved significantly with increasing years of medical school whereas Hren et al. (22) found that the highest level of positive attitude was observed among third-year students and those entering medical school. The likely explanation provided was that third-year students had completed the preclinical core curriculum. Additionally, students entering medical school directly from high school were noted to possess a predisposition toward a scientific approach to studying nature (22). Majority (27%) of the participants preferred the second professional year followed by the first professional year for starting their research training and the least preference was for the final year of medical school. A possible reason for this could be the lengthy academic syllabus in final-year which deters them from participating in research actively.

In the present study the primary motivation for engaging in research was their own interest followed by a desire to enhance their academics or to attain publications whereas peer pressure was the least motivating factor to engage in research which are similar to findings as reported in a study conducted by Pallamparty et al. (20). This was in contrast to previously conducted studies by Ahmed et al., Ibrahim et al., Siemens et al. and Griffin et al., where the majority of the participants were involved in research to facilitate their admission into postgraduate residency programs or career progression (12,14,17,24). This is due to the vastly different criteria for acceptance into a residency program in India from other nations where these studies were conducted. India facilitates admission into residency through an eligibility-cum-ranking examination (NEET PG) of multiple-choice question-based competitive exam conducted annually (25) whereas countries like Saudi Arabia (12) and Canada (17) where the studies have been conducted lay importance to curriculum vitae (CV), which includes research and publications as a criterion for acceptance into residency.

It is interesting to note that while the knowledge of research increased as the professional years advanced, the attitude towards research was found to be more positive in participants from pre- and para-clinical years in our study. Therefore, to bridge this gap between knowledge and attitude, we feel that medical students should be introduced to practicing research skills in their earlier years of medical school. This is further evident through our findings in our study where the maximum participants felt that research training should be started from the 2nd professional year of medical school.

While most of the participants generally had a positive attitude towards research, only 17% (68) had been a part of a research project, while only 4.72% (19) of the subjects had a publication. Poor level of participation in research was noted in previously conducted studies by Abushouk et al. and Amin et al. where only 24% and 17% subjects had participated in research (3,19). However, a study in Britain revealed encouraging results, with 49% of the medical students having taken part in an audit or a research project with 14% having submitted an article for publication. A reason for this encouraging result is that 51% engage in research for career progression since the British National Foundation training program awards points for publications and publication status to assess for specialist training posts. (24) In contrast to our findings, another study in India revealed that 34.3% of students have engaged in research with 17.4% having published in journals (26).

We found that the primary barriers among participants towards conducting research were lack of funds/laboratory equipment/infrastructure, lack of exposure of opportunities for research in the MBBS curriculum, lack of time, difficulty in finding a supervisor, lack of incentives for conducting research, and lack of knowledge and skills to conduct research. The majority of the studies have shown a lack of time as the major barrier that deters medical students from pursuing research (3,14,17,19,20,27). Similar to the findings in our study, the other major barriers to research include lack of training, skills, and knowledge (13,14,17), difficulty in finding a mentor (3,12,13,14,19), and lack of research facilities and infrastructure (3,13). A few barriers like lack of funds, laboratory equipment, and infrastructure may show regional differences as in developed nations like the USA where a study revealed that only 22% felt a scarcity of financial resources as a barrier towards research (27).

Medical students in India are introduced to basic concepts of research like formulating a research question for a study, the methods of collection, classification, analysis interpretation, and presentation of statistical data, and the principles and concepts of epidemiology and epidemiological study designs as a part of their core curriculum and are evaluated for the same during their 3rd professional year of training (28). However, the practical aspects of research like literature review, searching for a research question, protocol writing, etc. are still lacking in the curriculum. In an effort to correct this the undergraduate medical education board (UGMEB) updated its medical curriculum for undergraduates by introducing competency-based medical education (CBME) where two months were designated for elective rotations completion of the examination at the end of the third year (29). This two-month duration is split into two blocks, each lasting for 4 weeks. During block 1, participants will engage in activities either in a pre-selected preclinical, para-clinical, or other basic sciences laboratory, or work alongside a researcher involved in an ongoing research project and block 2 shall be done in a clinical department in the institution or as a supervised learning experience at a rural or urban community clinic. Previous studies have demonstrated that participation in an elective research experience has a positive impact across all personal and career characteristics (30). Furthermore, to promote research, the Indian council of medical research (ICMR) has introduced a short-term studentship program where undergraduates are encouraged to undertake research in their fields of interest by providing them with incentives and a certificate as a token of appreciation (31). Hence, we feel that this is a step in the right direction of generating future evidence-based medicine practitioners, who are not only adept at their clinical skills but are also involved in the development of newer diagnostic and treatment modalities through their active participation in research activities. However, the road towards achieving this goal is a long one and much improvement is needed, especially in developing countries like India. In a 2014 study by Ray et al. examining research output from Indian medical institutions, the findings underscored the concerning state of Indian medical research. The study revealed that 57% of the institutions had no publications included in the Scopus database. Additionally, a mere 4% of the institutes were responsible for generating over 100 papers annually (6). While the National Medical Commission (NMC) has mandated writing a thesis as a part of the medical postgraduate curriculum in India (32). However, a study in India by Dhaliwal et al. has revealed that only 30% of the theses were published in PubMed-indexed journals (33). A higher proportion of unpublished theses results in them being inaccessible to the majority of the scientific community and also raises suspicions about the quality of research thereby compromising the quality of postgraduate medical education.

To tackle this problem, we feel that research should be introduced in the curriculum from early on in the medical school years. Mandating research projects as a part of the curriculum has been shown to improve the research skills, literature search, and data analysis of the students (34). Establishing student research societies, organizing conferences, workshops, and journal club meetings, faculty-based mentorship programs, and providing incentives can also serve to increase the quality and quantity of medical research. Furthermore, it is vital that active measures be taken to develop the necessary infrastructure for performing medical research in an environment where research tends to be sidelined due to the overwhelming clinical burden that takes priority.

The limitation of our study is that the data was collected from a single medical school and cannot be generalized to a larger population. Given the cross-sectional nature of the study, it is not possible to establish a temporal relationship between the lack of knowledge and the identified barriers. Therefore, we advocate for the implementation of a multi-centre longitudinal or cohort study to effectively address this issue.

## Conclusions

Our study revealed that while most of the students had adequate knowledge and a positive attitude towards research, a notable deficiency in research participation was observed among the students. Lack of funds/laboratory equipment/infrastructure, lack of exposure to opportunities for research in the medical (MBBS) curriculum, and lack of time are few barriers to research. These challenges can be overcome by incorporating research as a part of the medical school curriculum from early years on, setting aside separate time for research, encouraging research electives, providing incentives to students engaging in research, developing appropriate infrastructure for conducting research, and establishing student research societies that can actively promote research through workshops, competitions, and mentorship (peer-based and faculty-based) programs.

## Data Availability

All data produced in the present study are available upon reasonable request to the authors.

## Acknowledgement

We would like to thank all the students who have participated in the study.

## Ethics approval and consent to participate

Ethical approval was taken from Institutional Ethical Committee (IEC) with IEC no. F.no. 5(2)/2023/BSAH/DNBCommittee 3518-3519. Written informed consent was taken from all the participants.

## References

1. World Health Organization. Regional Office for the Western Pacific. (2001). Health research methodology: a guide for training in research methods. 2nd ed. WHO Regional Office for the Western Pacific. Available from: https://apps.who.int/iris/handle/10665/206929 [Last accessed June 19, 2023]

2. Vernekar P, Kunkolienkar R, Fernandes R, Chiranth AB, Khandeparkar V, Cacodcar JA. Perceptions, attitudes, practices and barriers towards research amongst postgraduate medical students in Goa. International Journal of Contemporary Medical Research 2020;7(9): 11–I6.

3. Abushouk AI, Hatata AN, Omran IM, Youniss MM, Elmansy KF, Meawad AG. Attitudes and perceived barriers among medical students towards clinical research: A cross sectional study in an Egyptian medical school. J Biomed Educ. 2016 Oct;2016(7):5490575.

4. Straus SE, Straus C, Tzanetos K, Under the auspices of the International Campaign to Revitalise Academic Medicine. Career choice in academic medicine: systematic review. Journal of general internal medicine. 2006 Dec;21(12):1222–9.

5. Academic Research and Development, Science & Engineering Indicators 2018, National Science Board, 2018. Arlington, VA: National Science Foundation Available from:https://www.nsf.gov/statistics/2018/nsb20181/report/sections/academic-research-anddevelopment/highlights [Last accessed May 19, 2022]

6. Ray S, Shah I, Nundy S. The research output from Indian medical institutions between 2005 and 2014. Current Medicine Research and Practice. 2016 Mar 1;6(2):49–58.

7. Sharma SK, Thatikonda N, Ukey UU. Knowledge, Attitude, Practice and Barriers for Research amongst Medical Students of GMC, Nagpur. J Res Med Dent Sci, 2021, 9 (4):41–7.

8. Garg R, Goyal S, Singh K. Lack of research amongst undergraduate medical students in India: it’s time to act and act now. Indian pediatrics. 2017 May;54(5):357–60.

9. Juhi A. Approach toward research among the medical undergraduates studying in Apollo Medical College, Hyderabad. National Journal of Physiology, Pharmacy and Pharmacology. 2020;10(9):749–54.

10. Pourhoseingholi MA, Vahedi M, Rahimzadeh M. Sample size calculation in medical studies. Gastroenterol Hepatol from bed to bench. 2013;6(1):14–7

11. Omprakash A, Kumar AP, Ramaswamy P, Sathiyasekaran BW, Ravinder T. Assessment of Knowledge, Attitude, Perceived Barriers towards Research among First Year Undergraduate Medical Students: A Study from Chennai, Tamil Nadu, India. Journal of Clinical & Diagnostic Research. 2019 Nov 1;13(11).

12. Ahmed NJ. Evaluation of Medical Sciences Studentsâ€™ Perception, Barriers, and Attitude toward Scientific Research: A Cross-sectional Study in Makkah Region, Saudi Arabia. Asian Journal of Pharmaceutics (AJP). 2021 Nov 1;15(3).

13. Vairamani CR, Akoijam BS. Knowledge, attitude and perceived barriers towards conducting research among students in a medical college, India. Int J Community Med Public Health. 2018 Jun;5(2):806–10.

14. Ibrahim NK, Fetyani DM, Bashwari J. Assessment of the research-oriented knowledge, attitude and practice of medical students and interns of the King Abdulaziz University, Jeddah and the adoption of a research-intervention educational program. Rawal Med J. 2013 Oct;38(4):432–9.

15. Ismail IM, Bazli MY, O’Flynn S. Study on medical student’s attitude towards research activities between University College Cork and Universiti Sains Malaysia. Procedia-Social and Behavioral Sciences. 2014 Feb 21;116:2645–9.

16. Kumar HH, Jayaram S, Kumar GS, Vinita J, Rohit S, Satish M, et al. Perception, practices towards research and predictors of research career among UG medical students from coastal South India: A cross-sectional study. Indian Journal of Community Medicine: Official Publication of Indian Association of Preventive & Social Medicine. 2009 Oct;34(4):306.

17. Siemens DR, Punnen S, Wong J, Kanji N. A survey on the attitudes towards research in medical school. BMC medical education. 2010 Dec;10(1):1–7.

18. Meraj L, Gul N, Zubaidazain IA, Iram F, Khan AS. Perceptions and attitudes towards research amongst medical students at Shifa College of Medicine. JPMA. 2016;66(2):165–9.

19. Amin TT, Kaliyadan F, Al Qattan EA, Al Majed MH, Al Khanjaf HS, Mirza M. Knowledge, attitudes and barriers related to participation of medical students in research in three Arab Universities. Educ Med J. 2012 Jun 1;4(1):47–55.

20. Pallamparthy S, Basavareddy A. Knowledge, attitude, practice, and barriers toward research among medical students: A cross-sectional questionnaire-based survey. Perspectives in clinical research. 2019 Apr;10(2):73.

21. Khan H, Khawaja MR, Waheed A, Rauf MA, Fatmi Z. Knowledge and attitudes about health research amongst a group of Pakistani medical students. BMC medical education. 2006 Dec;6(1):1–7.

22. Hren D, Lukić IK, Marušić A, Vodopivec I, Vujaklija A, Hrabak M, et al. Teaching research methodology in medical schools: students’ attitudes towards and knowledge about science. Medical education. 2004 Jan;38(1):81–6.

23. Vujaklija A, Hren D, Sambunjak D, Vodopivec I, Ivaniš A, Marušić A, et al. Can teaching research methodology influence students’ attitude toward science? Cohort study and nonrandomized trial in a single medical school. Journal of Investigative Medicine. 2010 Feb;58(2):282–6.

24. Griffin MF, Hindocha S. Publication practices of medical students at British medical schools: experience, attitudes and barriers to publish. Medical teacher. 2011 Jan 1;33(1):e1–8.

25. National Board of Examinations in Medical Sciences (NBEMS). ENTRANCE EXAMINATION-NEET PG. Available from: https://natboard.edu.in/viewnbeexam?exam=neetpg. (Last accessed: November 28, 2023).

26. Chellaiyan VG, Manoharan A, Jasmine M, Liaquathali F. Medical research: Perception and barriers to its practice among medical school students of Chennai. Journal of education and health promotion. 2019;8.

27. Ho A, Auerbach A, Faulkner JJ, Guru SK, Lee A, Manna D. Barriers to research opportunities among osteopathic medical students. Journal of Osteopathic Medicine. 2023 Feb 1;123(4):187–94.

28. Medical Council of India. Competency based Undergraduate curriculum for the Indian Medical Graduate, 2018. Vol. 2; pg. 44-46. Available from: https://www.nmc.org.in/wp-content/uploads/2020/01/UG-Curriculum-Vol-II.pdf. [Last accessed: 23 Nov, 2023]

29. Medical Council of India. Electives for the Undergraduate Medical Education Training Program, 2020: p 1-30. Available from: https://www.nmc.org.in/wp-content/uploads/2020/08/Electives-Module-20-05-2020.pdf [Last accessed: 23 Nov, 2023]

30. Cuschieri A, Cuschieri S. Analysing the impact of an elective research experience on medical students’ research perceptions. Medical Science Educator. 2023 Feb;33(1):157–64.

31. Short Term Studentship (STS). Indian Council of Medical Research (ICMR). Government of India. Available from: https://main.icmr.nic.in/content/short-term-studentship-sts [Last accessed: 23 Nov, 2023]

32. National Medical Commission (NMC). P.G. (Postgraduate) Medical Education Regulations, 2000. Available from: https://www.nmc.org.in/rules-regulations/p-g-medical-education-regulations-2000/ [Last accessed: 23 Nov, 2023]

33. Dhaliwal U, Singh N, Bhatia A. Masters theses from a university medical college: publication in indexed scientific journals. Indian journal of ophthalmology. 2010 Mar;58(2):101.

34. Kaur R, Hakim J, Jeremy R, Coorey G, Kalman E, Jenkin R, et al. Students’ perceived research skills development and satisfaction after completion of a mandatory research project: results from five cohorts of the Sydney medical program. BMC Medical Education. 2023 Jul 12;23(1):502.

